# Onset, duration, and persistence of taste and smell changes and other COVID-19 symptoms: longitudinal study in Israeli patients

**DOI:** 10.1101/2020.09.25.20201343

**Authors:** Hadar Klein, Kim Asseo, Noam Karni, Yuval Benjamini, Ran Nir-Paz, Mordechai Muszkat, Sarah Israel, Masha Y. Niv

## Abstract

**Objectives:** The multifaceted manifestation of COVID-19 requires longitudinal characterization of symptoms, to aid with screening and disease management.

**Methods:** Phone interviews and follow-ups were completed with 112 mostly mild COVID-19 RT-PCR-positive adult patients, over a six-months period.

**Results:** More than one symptom at disease onset was experienced by ∼70% of the patients. About 40% of the patients experienced fever, dry cough, headache, or muscle ache as the first symptom. Fatigue, if reported, usually was the first to appear. Smell and taste changes were experienced 3.9 ± 5.4 and 4.6 ± 5.7 days (mean ± SD) after disease onset and emerged as first symptom in 15% and 18% of patients, respectively. Fever had the shortest duration (5.8 ± 8.6 days), and taste and smell changes were the longest-lasting symptoms (17.2 ± 17.6 and 18.9 ± 19.7 days, durations censored at 60 days). Longer smell recovery correlated with smell change severity. Cough, taste change and smell change persisted after negative RT-PCR tests (in 20%, 26% and 29% of the patients in total). At six-months follow-up, 46% of the patients had at least one unresolved symptom, most commonly fatigue (21%), chemosensory changes (14%) or breath difficulty (9%).

**Conclusions:** More than one symptom typically occurred at disease onset. Chemosensory changes and cough persisted after negative RT-PCR in a quarter of the patients. Almost half of the patients reported at least one unresolved symptom at six-months follow up, mainly fatigue, smell changes and breath difficulty. Our findings highlight the prevalence of long-lasting effects of COVID-19.

## BACKGROUND

The coronavirus disease (COVID-19) has become a worldwide pandemic, with more than 37 million cases and over a million deaths (World Health Organization, October 12^th^, 2020). COVID-19 symptoms, severity and duration vary widely[1–4]. Researchers and patient groups are urging clinicians and scientists to use the term “long COVID” and to listen to patients voices[5].

COVID-19 initially targets the human respiratory system, with the most common symptoms according to the Centers for Disease Control and Prevention (CDC) being fever, cough, shortness of breath, fatigue, muscle aches, headache, new loss of smell or taste, sore throat, congestion, nausea/vomiting and diarrhea. Other commonly reported symptoms also include sputum production, hemoptysis and lack of appetite[6,7]. The symptoms order of appearance, severity, durations and persistence after recovery, is only partly studied[8–10] and is the focus of our study here.

## METHODS

### Aim and setting

This is a follow-up study of patients that participated in our previous study, see Karni and Klein et al.[11], performed in Israel, from April 2020 to October 2020. The study comprised of Israeli residents aged ≥18 years with positive COVID-19 RT-PCR results, who were recruited via social media (Twitter and Facebook) and word of mouth for phone interviews. We excluded severely ill patients, and non-Israeli residents. Participants were not screened or targeted for experiencing chemosensory changes. Informed consent was obtained from all participants. The study was approved by the Hadassah Medical Center Helsinki Committee (permit number 0236-20-HMO). Upon agreement to participate, the explanation of the interview, including questions on taste and smell, were read to the participants.

### General design

The questionnaire was developed in parallel with the Global Consortium for Chemosensory Research, GCCR[12]. The questionnaire[11] had five parts: 1) General information (e.g., age, gender); 2) Medical history (e.g., medical conditions, chronic medications use); 3) Current illness (e.g., 23 physical signs and symptoms, RT-PCR swab test results and dates, subjective recovery feeling); 4 and 5) Smell and taste: Participants were instructed to rate their sense of smell/taste before, during and after their illness, on a scale from 1-10 (1 corresponding to “no sense of smell” and 10 to excellent sense of smell). Data was kept in Compusense Cloud on-line software (Compusense Inc., Guelph, ON, Canada).

### Patients follow-up

COVID-19 positive patients from our previous Karni and Klein et al. study[11], were followed here to monitor the progress of their illness and recovery. Out of 144 RT-PCR-positive patients, 114 had answered the phone, of which two had missing information regarding rating of chemosensory symptoms, resulting in 112 patients answering the first questionnaire. Recovered patients’ category refers to those who had recuperated from all their symptoms and received two consecutive negative RT-PCR test results, with one or more days apart. Patients who either did not recover from their symptoms in the first questionnaire, or did not get two consecutive negative RT-PCR results at the time of answering the first questionnaire, were contacted 3 weeks after the initial interview, for their second interview (n=101). If they had not fully recovered at their first follow-up, they were contacted again 3 weeks after their second interview, for their third interview (n=46). Additionally, all patients were contacted again approximately six months after their fourth interview (n=105, 7 patients did not answer the phone). Data from these four interviews was analyzed for symptoms characteristics: onset, duration, persistence post recovery and severity.

### Data analysis

The participants were asked about 27 symptoms (see web-only Supplementary Figure S1). 7 symptoms occurred in more than 50% of the patients (taste change, smell change, fever, dry cough, muscle aches, headache, lack of appetite) and were therefore included in the analysis. All but lack of appetite are official CDC symptoms as of October 2020. 7 additional official CDC symptoms were included as well: productive cough, shortness of breath, fatigue, sore throat, congestion, nausea/vomiting, and diarrhea. “Fatigue” did not explicitly appear in the questionnaire but was self-reported in 18% of patients under “any other symptoms” and was therefore included in the final analysis.

*Disease onset* is defined as the first appearance of any of the COVID-19 symptoms listed above.

*“Number of days after disease onset”* was the date of symptoms’ appearance minus the date of the first symptom to occur. Mean ± SD number of days since illness started was then calculated for each symptom.

*Persistent* symptoms are defined as symptoms that persist after the patient’s recovery as described above.

*Unresolved* symptoms refer to those that were unresolved at the six-months follow-up. Order of appearance were calculated from the onset dates reported by the participants for each symptom. Duration was calculated from the onset and resolution date of each symptom. In case patients reported unresolved symptoms or could not report a precise resolution date between the 6-weeks follow-up to the six-months follow-up, their symptoms’ duration was censored at 60 days, the longest reported duration by the 6-weeks follow-up. This resulted in 10% of the data being censored.

Maximal severity of smell (taste) change was calculated as the difference between the rating of the respective sense level before the illness and its rating during the illness, rated on a 1-10 scale.

### Statistical analysis

Analyses were conducted using the statistical software R (https://www.r-project.org/). Descriptive statistics are presented as the mean and standard deviation.

Correlations were measured using Pearson correlation coefficients, and p-values are based on correlation t-tests. A level of P<0.05 was used to determine statistical significance. P-values for the pair-wise comparisons between symptom durations were calculated using a two-sided Wilcoxon rank-sum test, and adjusted for multiplicity using the BH procedure[13]. A level of P<0.05 for the adjusted p-values was used to determine statistical significance.

## RESULTS

### Patients’ characteristics

Completed questionnaires were obtained from 112 COVID-19 positive patients. The median age of the respondents was 35 ± 12 years (mean ± SD), 72 men. 6 patients were hospitalized (received respiratory support during their hospitalization and / or were hospitalized in the intensive care unit) and the remaining 106 were classified as ambulatory patients.

### Order of symptoms’ appearance

Symptoms usually appeared concomitantly: The first and second symptoms to appear were accompanied by additional symptoms in 69% and 45% of the patients, respectively. Headache, fever, dry cough, and muscle aches were the first to appear in over a third of patients, at 2.0 ± 5.0, 1.8 ± 4.6, 1.4 ± 3.4 and 2.1 ± 5.1 days (mean ± SD), respectively, after disease onset. Smell and taste changes appeared later, at 3.9 ± 5.5 and 4.6 ± 5.7 days (mean ± SD), respectively, after disease onset (Figure 1B) and were the first symptoms in only 15% and 18% of patients. Fatigue was a first symptom in 14% of patients and was the first symptom in the majority of patients (80%) in which it occurred (Figure 1A). Each of the 14 symptoms analyzed here could occur as first or co-first. Vomiting, breathing difficulty and diarrhea were the most spread out in the order in which they appear (see web-only Supplementary Figure S2).

**Figure 1.**
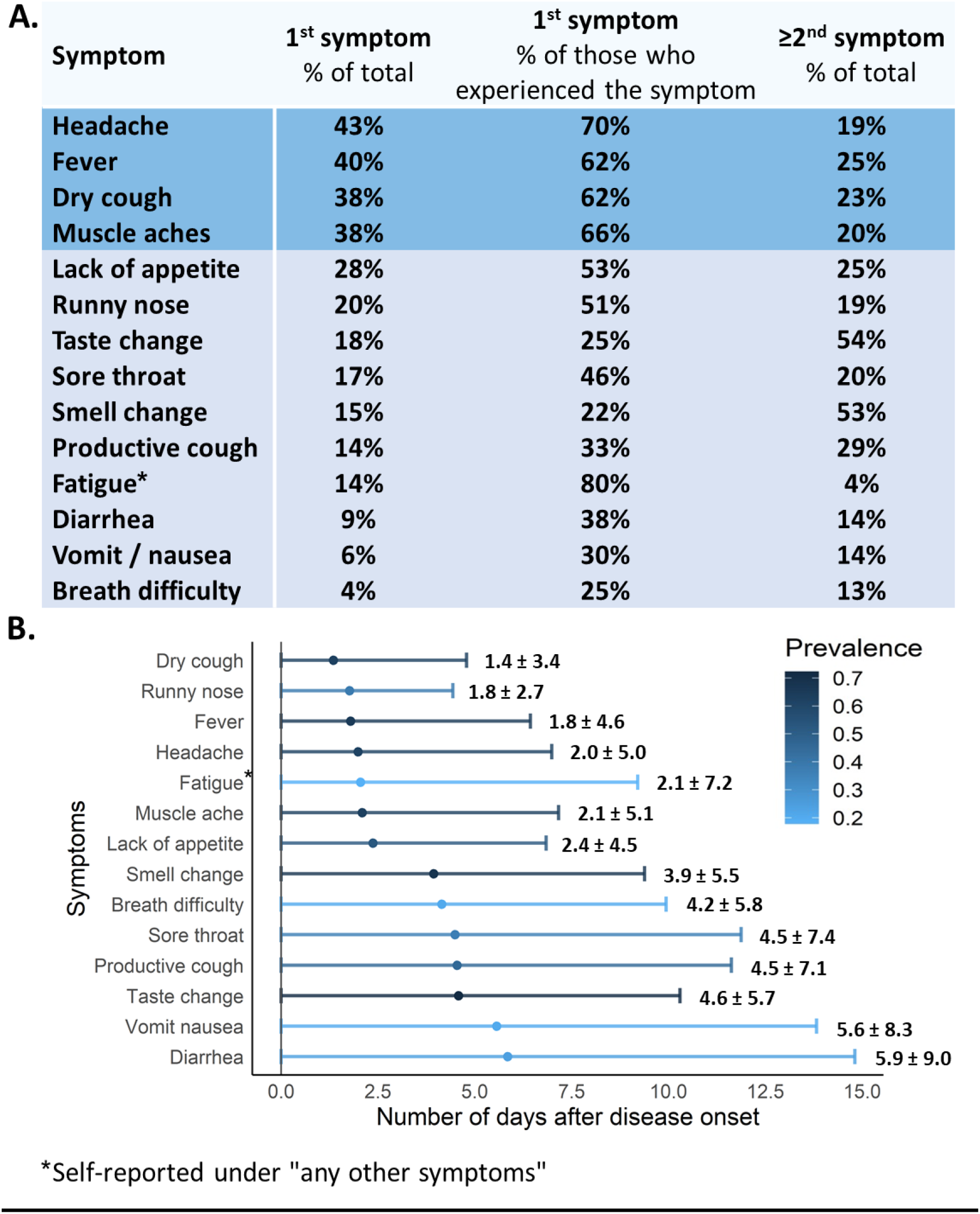
Symptoms order. **A**. Prevalence of patients for which symptoms appeared as 1^st^ and ≥2^nd^ symptoms. Symptoms that appeared as 1^st^ in more the third of the patients are presented with darker blue background, lower prevalence is presented in lighter blue background. **B**. Y axis lists symptoms and X axis shows number of days after disease onset. The mean ± SD number of days after disease onset is presented for each symptom. Coloring is according to prevalence, with darkest color indicating most prevalent symptoms.

Vomiting and diarrhea appeared after the highest number of days since disease onset (5.6 ± 8.3 and 5.9 ± 10.0, respectively) (Figure 1B).

### Distribution of symptoms durations and persistence

Figure 2 depicts the durations of the most prevalent symptoms (>50%) among 112 COVID-19 patients. Fever had shorter duration than all other symptoms (5.8 ± 8.6 (mean ± SD)) (P<0.05). Muscle aches, lack of appetite and headache (6.9 ± 8.1, 7.8 ± 6.9 and 7.8 ± 8.5 (mean ± SD)) had shorter mean of censored durations than those of dry cough and changes of taste and smell (14.8 ± 14.6, 17.2 ± 17.6 and 18.9 ± 19.7 (mean ± SD)) (P<0.05). In these patients, the most prolonged symptom was breath difficulty, in some unresolved even after 213 days.

**Figure 2.**
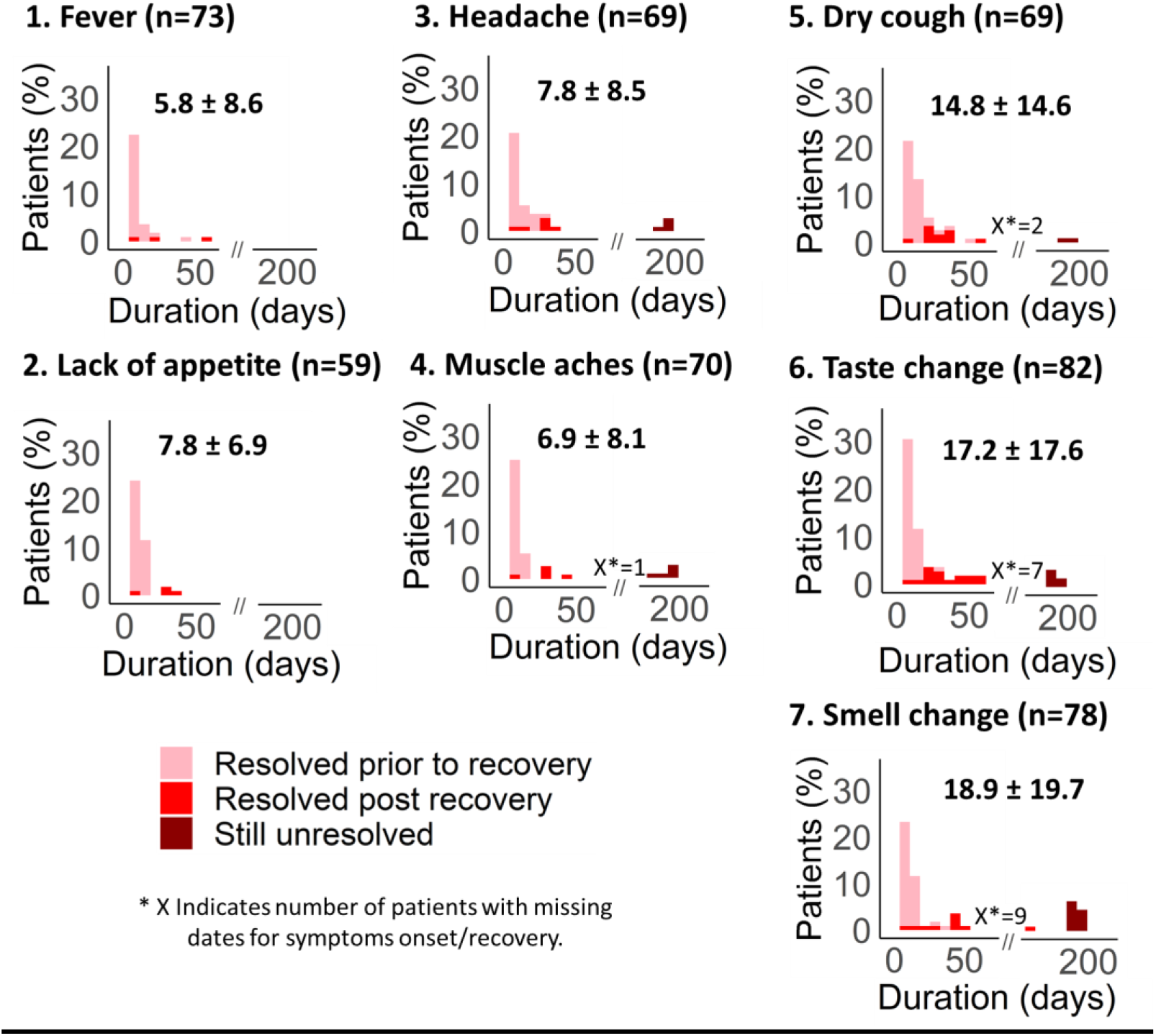
Symptoms durations. Duration pattern graphs are presented for each symptom and colored according to symptom’s persistence after recovery (resolved prior to recovery are colored pink, resolved post recovery are colored red and unresolved symptoms at six-months follow-up are colored dark red). Y axis shows % of patients and X axis shows duration of symptoms (days). X represents the number of patients with missing dates for symptoms onset/recovery. Number of patients and the mean ± SD of symptoms censored durations (days) are presented for each graph.

Patients with smell change, taste change, and dry or productive cough still had these symptoms post recovery (41%, 35%, 28%, respectively, equivalent to 29%, 26% and 20% of all patients) (Figure 2). Persistent taste change within 6-weeks was usually accompanied by a persistent smell change in the same patient, and in some cases also accompanied by a persistent cough (productive or dry). In 14% of the patients with cough, it persisted without persistent chemosensory change (Figure 3A).

**Figure 3.**
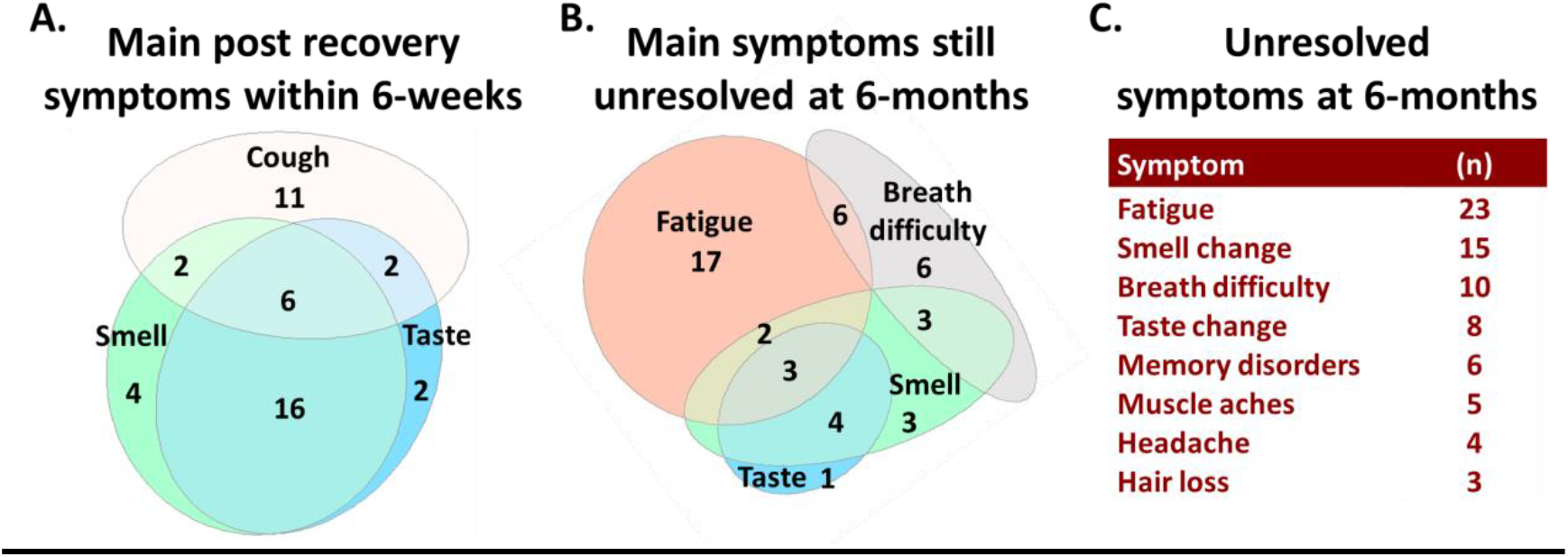
Symptoms persistence. **A**. Euler-diagram of the main persistent symptoms post recovery collected during 6-weeks of follow-ups. The number of patients experiencing a symptom as persistent is marked for each circle. **B**. Euler-diagram of the main unresolved symptoms collected at the six months follow-up. The number of patients experiencing a symptom as unresolved is marked for each circle. **C**. Table of unresolved symptoms at six-months follow-up and number of patients experiencing them.

At six-months follow-ups, 51 patients (46%) still reported unresolved symptoms, mainly fatigue (21%), chemosensory changes (14%) and breath difficulty (9%). Fatigue, breath difficulty, memory disorders and hair loss, were not typically reported during the 6-weeks follow-ups (thus “new symptoms”), while other symptoms such as muscle aches, headache and chemosensory changes usually carried over from previous interviews (Figure 3C). For additional symptoms at six-months follow-up, see web-only Supplementary Table S1).

Of the 23 patients that had not fully recovered during 6-weeks of follow-ups, 17 reported smell changes. 11 of these patients still reported smell changes at six-months. 4 patients who reported they recovered from their smell change at the 6-weeks follow-up, stated at the six-months follow-up that their sense of smell continued fluctuating and remained unresolved. Patients who experienced chemosensory changes at six-months follow-up had either smell changes only (n=8), or smell and taste changes (n=7). Seven patients reported distorted sense of smell.

### Smell change severity and recovery

The durations of changes in smell and taste reported at the 6-weeks follow-up, were highly correlated (0.82, p<0.001) (Figure 4A).

**Figure 4.**
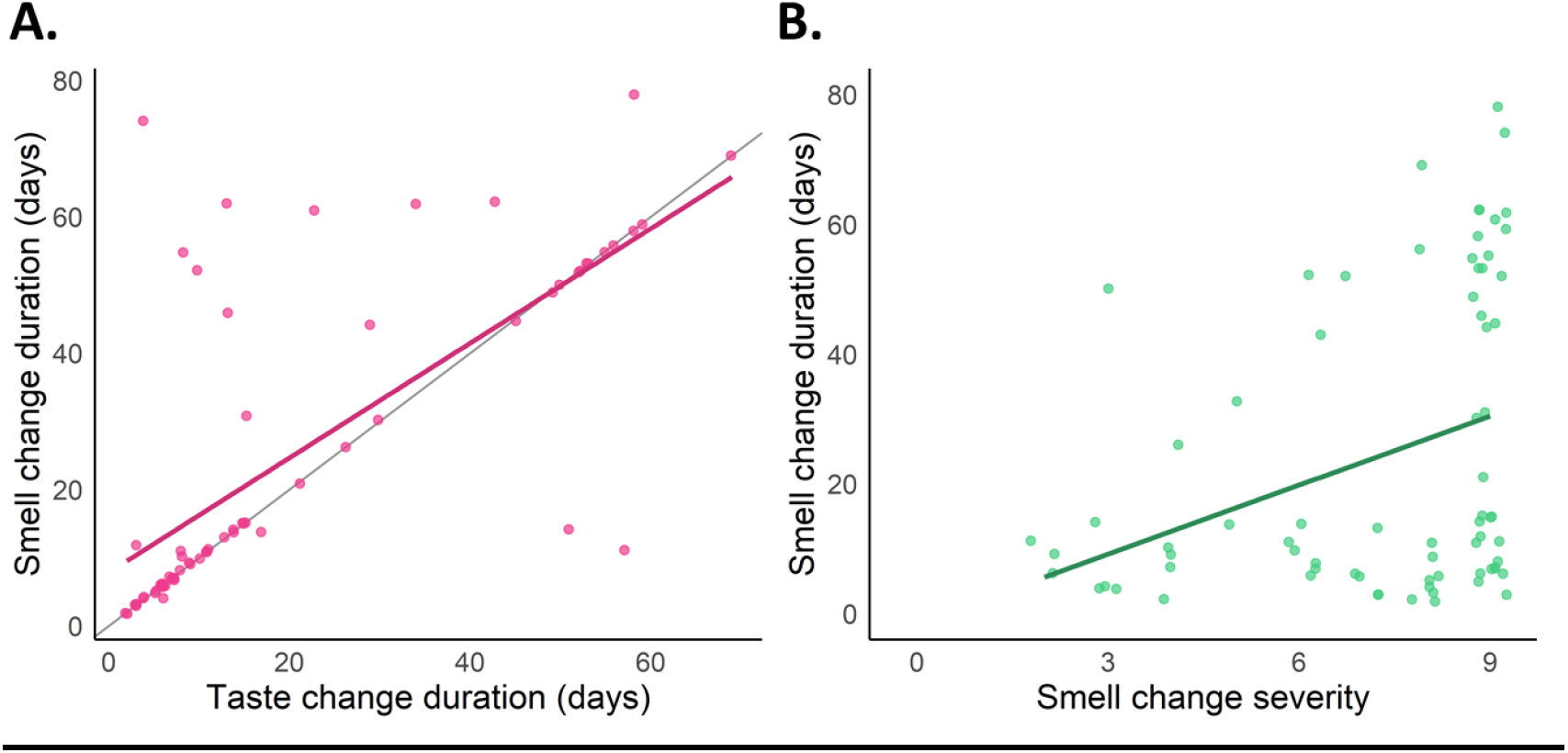
Correlations of smell and taste change characteristics. **A**. Correlation of smell and taste change durations (n=67). X axis shows taste change duration and Y axis shows smell change duration (days). The diagonal grey line shows optimal correlation of 1 and the pink line shows the calculated correlation of 0.82. **B**. Correlations between smell change severity and its duration (n=76). X axis shows smell change severity and Y axis shows smell change duration (days). Green line shows the calculated correlation of 0.34.

As seen in Figure 4B, a weak yet significant correlation was found between smell change severity and its duration (correlation 0.34, p=0.003). However, there was no correlation between taste change severity and duration (see web-only Supplementary Figure S3). Moreover, it is apparent that some patients’ smell change had recovered independently of how severe it was, while for another group of patients, it was closely dependent on its severity (Figure 4B). No special characteristics of subpopulations could be identified.

Chemosensory recovery patterns indicate full recovery in some patients and a slower and gradual recovery of the senses in others (see web-only Supplementary Figure S4).

## DISCUSSION

Smell and taste changes had emerged as distinctive COVID-19 symptoms[11,14]. Additional symptoms are emerging for long haulers[5].

Our study reveals that smell and taste were the first symptoms to appear, in 15% and 18% of cases, respectively, either with or without other symptoms. Spinato et al.[15] had found that the onset of smell or taste changes was prior to other symptoms in 11.9% of cases and concomitant with other symptoms in 22.8% of cases. Our slightly lower prevalence as first symptoms, could be due to recall bias caused by retrospective data collection.

We found the means of censored durations of taste change to be 17.2 ± 17.6 days and of smell change to be 18.9 ± 19.7 days (mean ± SD), and their post-recovery persistence to be prevalent in 35% and 41% of patients (usually by the same patient). Additionally, cough, either dry or productive, was also found to persist in approximately 27% of patients who reported experiencing this symptom.

Different reports found that the time for chemosensory recovery is ranging from 8 ± 6.4 to 11.6 ± 6.2 days (mean ± SD)[16,17], others found their persistence post recovery in the range of 20% to 78% of patients[10,18]. The higher numbers in these studies could be due to smaller samples or different patients’ recruitment.

Our long (up to six-months) follow-up enabled to capture longer duration of chemosensory changes than those reported by others[16,17,19]. Moreover, our detailed phone interviews provide reliable data which is not readily available in online studies.

Importantly, we monitored quantitative recovery back to normal levels, possibly accounting for the longer recovery rate we found for the chemosensory changes and higher persistence post-recovery than when binary classification was used[19].

We found smell (but not taste) change severity to be correlated with its time to recover. However, while for a group of the patients, the time necessary for their smell change recovery correlated with magnitude of the change they had experienced, for another group, the severity of their smell change was unrelated to the time it took to recover. Such grouping was also reported by Gerkin et al.[14], but characteristics that might account for the differences between these two groups could not yet be unraveled and require further exploration with large groups of patients.

Concerning other COVID-19 symptoms, we found that even though their order of appearance is varied, headache, fever, dry cough, and muscle aches were often (38% −43%) the first symptoms to appear. Additionally, out of the 18% patients who experienced fatigue, 80% had experienced it as a first symptom (14% of all patients). Of note, the prevalence of fatigue in our study was lower than previously reported 36%-46%[20,21], probably because here it was not explicitly part of the questionnaire, but rather self-reported under the “any other symptom” question.

Overall, 51 patients had not fully recovered from their symptoms six months after their first questionnaire, with most of them reporting unresolved fatigue, chemosensory changes, and breath difficulty. These unresolved symptoms can occur together or separately, and some of them were not reported during the initial 6-weeks follow-up.

### Study limitations

Our sample did not include severely ill patients, and therefore is relevant for light to moderately ill patients only. No objective testing was performed, and the information was self-reported by the participants. Additionally, the retrospective data collection method used in this study may have caused recall bias.

Patients were contacted for the second and third interviews only if they have not reported recovery. In principle, it is possible that some of the recovered patients could have recurrences of symptoms as well[22]. Therefore, for the six-months follow-up we recontacted all study participants for reports of unresolved symptoms. Long period studies of larger groups of COVID-19 patients, with more follow-ups are needed to further monitor and characterize manifestations of “long COVID”[23].

## Conclusions

We carefully mapped the start and end dates of various COVID-19 symptoms of 112 mild patients in Israel. The results indicate that despite large variability in symptoms onset and duration, some symptoms (headache, fever, dry cough, and muscle aches) are common as first or co-first symptoms. Fatigue, if occurs, usually appears as a first symptom. Taste and smell changes are not typically first, and appear on average, 3.9 ± 5.4 and 4.6 ± 5.7 days (mean ± SD), respectively, after disease onset. Their recovery can be either fast, or longer and gradual, sometimes accompanied by cough. Interestingly, while dry cough appears early (1.4 ± 3.4 days (mean ± SD)) and productive cough later (4.5 ± 7.1 days (mean ± SD)), both types of cough may persist post recovery. At six-months follow-up, 46% of patients experienced unresolved symptoms, with symptoms either continuing throughout the illness (e.g., chemosensory changes), or appearing as new (e.g., memory disorders).

This information regarding their symptoms’ durations and persistence post recovery, paves the way to further long COVID studies and advances towards better understanding and management of the disease.

## Data Availability

The datasets used and/or analysed during the current study are available from the corresponding author on reasonable request.

## Authors’ contributions

NK and MYN initiated the research, SI, NK and MM wrote the IRB proposal, NK and HK recruited patients, NK, MYN, YB, SI and MM designed the research, SI, MM, RNP and MYN supervised the research, HK carried out the interviews, HK and MYN drafted the manuscript, KA and YB preformed the statistical analysis, KA and HK created figures and tables. All authors contributed to writing and approved the final manuscript.

## LIST OF ABBREVIATIONS

CDC: Centers for Disease Control and Prevention
SARS-CoV-2: Severe Acute Respiratory Syndrome Coronavirus 2
SD: Standard Deviation
GCCR: Global Consortium for Chemosensory Research
RT-PCR: Real Time Polymerase Chain Reaction
URI: Upper Respiratory tract Infections

## Ethics approval

The study was conducted in accordance with Helsinki committee and the required ethics approval was granted (reference number HMO-0236-20).

## Ethics, consent, and permissions

Written informed consents for publication of patients’ clinical details were obtained from the patients

## Acknowledgements

We thank the Global Consortium for Chemosensory Research (GCCR) team for fruitful collaborations and discussions. We thank Yehuda Tarnovsky for help with patient’s recruitment.

