## Supplementary material for "Onset, duration, and persistence of taste and smell changes and other COVID-19 symptoms: longitudinal study in Israeli patients"

| 27 symptoms: | | |
|---|---|---|
| ≥50% | <50% | |
| **Taste change** | **Productive cough** | Bad taste in mouth |
| **Smell change** | **Runny nose** | Voice change |
| **Fever** | **Sore throat** | Eyes burn |
| **Dry cough** | Chemesthesis change | Dizziness |
| **Muscle aches** | **Diarrhea** | Vision changes |
| **Headache** | Chest pain | Coated tongue |
| Lack of appetite | Abdominal pain | Ears pressure |
| | **Breath difficulty** | Lacrimation |
| | **Vomit/nausea** | Hearing change |
| | **Fatigue** | Eyes discharge |

**Figure S1. 27 symptoms of the questionnaire.** Symptoms with prevalence higher than 50% are presented with orange background, and those with prevalence lower than 50% are presents with light blue background. Official CDC symptoms are bolded. Symptoms that were included in the final analysis are underlined.

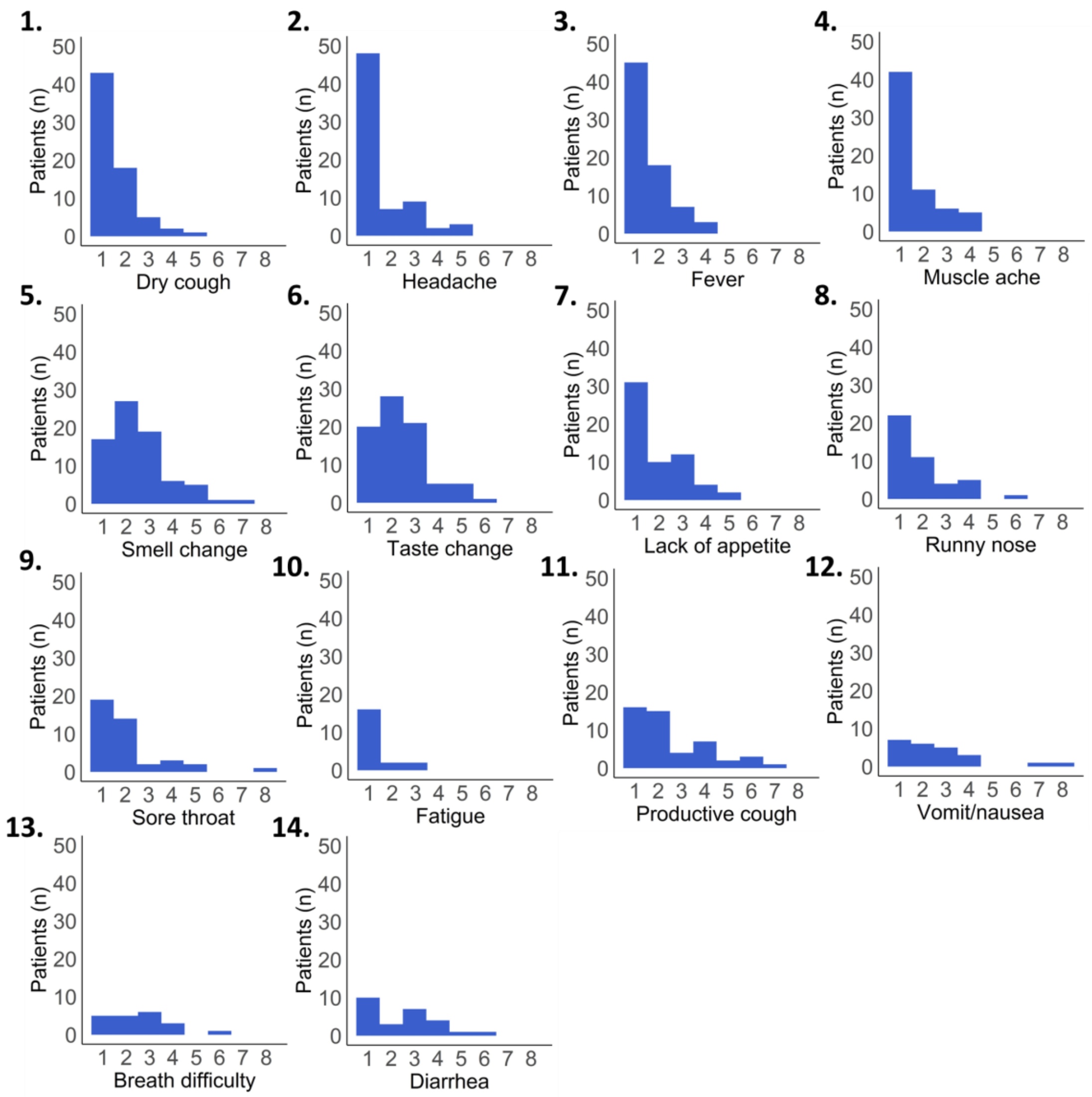


**Figure S2. Distributions of symptoms' order of appearance.** Y axis shows number of patients (n) and X-axis is order of appearance of the specified symptom (first symptom to appear or followed other symptoms).

**Table S1. Unresolved symptoms at 6-months follow-up.** Number of patients experiencing each symptom is listed.

| Symptom | Unresolved |
|---|---|
| Fatigue | 23 |
| Smell change | 15 |
| Breath difficulty | 10 |
| Taste change | 8 |
| Memory disorders | 6 |
| Muscle aches | 5 |
| Headache | 4 |
| Hair loss | 3 |
| Nose blockage | 2 |
| Legs pain | 2 |
| Eye disorders | 2 |
| Low physical performance | 2 |
| Vomit | 1 |
| Throat ache | 1 |
| Chest pain | 1 |
| Dizziness | 1 |
| Knee pain | 1 |
| Mouth sores | 1 |
| Hands pain | 1 |
| Ear pain | 1 |
| Paresthesia | 1 |
| Palpitations | 1 |
| Anemia | 1 |
| Anxiety | 1 |
| Dry cough | 1 |
| Hearing disorder | 1 |
| Rib pain | 1 |
| Depression | 1 |
| Concentration Disorders | 1 |
| Runny nose | 1 |
| Abdominal pain | 1 |
| Diarrhea | 1 |

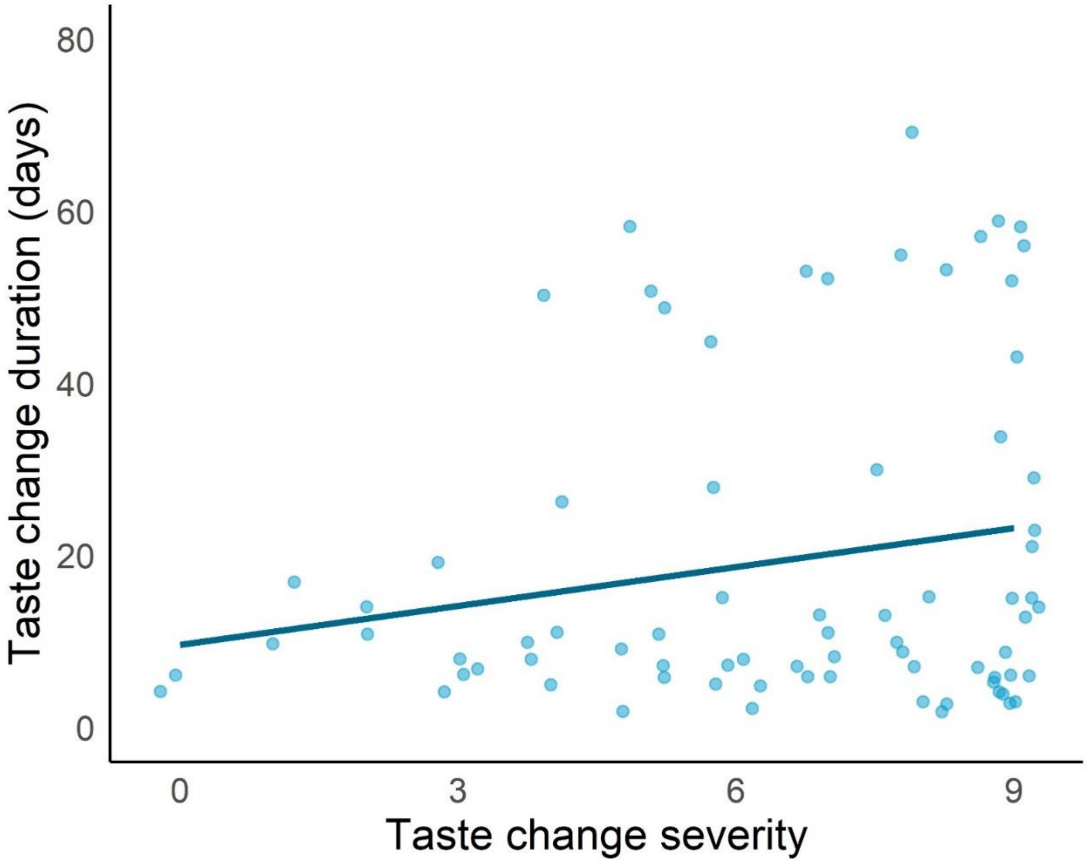


**Figure S3. Correlation between taste change severity and duration (n=76).** Y axis shows taste change duration (days) and X axis shows taste change severity.

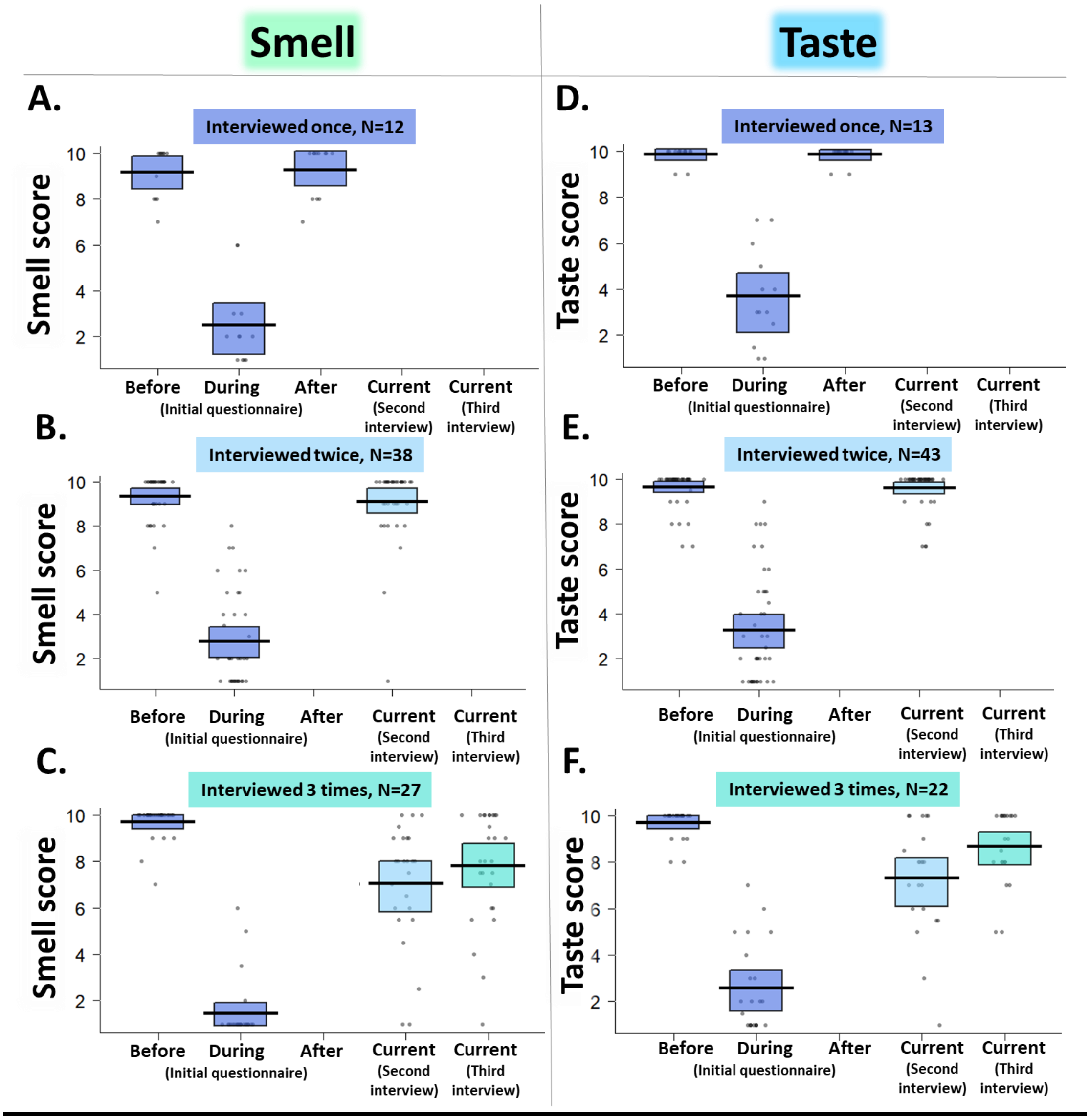


**Figure S4. Smell and taste ratings over 6-weeks follow-ups. A-C.** Smell scores of patients recovered at the initial **(A)**, second **(B)**, and third **(C)** interviews. **D-F.** Taste scores of patients recovered at the initial **(D)**, second **(E)**, and third **(F)** interviews. We plot the mean and the confidence interval for the mean. Y axis shows rating of smell/taste ability and X axis depicts 5 time points ratings: before, during and after their illness, rated at initial questionnaire (blue), and at two following questionnaires (3-weeks (light blue) and 6-weeks (turquoise) follow-ups).
